# Evaluating the Effectiveness of Point-of-Entry UV Treatment for Cistern Water Among Households in the US Virgin Islands

**DOI:** 10.1101/2022.10.18.22281228

**Authors:** Lee Voth-Gaeddert, Douglas Momberg, Kela Brathwaite, Andrew Schranck, Mandy Lemley, Stephen Libbey

## Abstract

UV water treatment can be a viable option for point-of-entry applications among households utilizing private water sources. In the US Virgin Islands (USVI), the primary water source is roof-harvested rainwater, collected in large cisterns and supplied to household taps via a pump. While diversification of water sources provides increased resilience to climate change, literature suggests rainwater catchment systems are at high risk of microbial contamination. One option USVI households have is UV systems. However, limited data is available on UV system effectiveness for USVI installations while these systems can be expensive. Therefore, Love City Strong, a local NGO, piloted a multi-year UV access program which included free UV systems with prefiltration along with installation and monthly household visits for up to 12 months including water quality testing. In addition, due to the significant costs associated with the prefiltration portion of the system, a pilot study was conducted to evaluate the effectiveness of the UV systems without prefiltration.

The results from the UV system access program demonstrated that *E. coli* was not detected in 95.2% of tap samples (n=271). Among samples with detectable levels of *E. coli* and total coliforms, turbidity was lower compared to samples with non-detections. Field teams reported user error was often identified in association with *E. coli* detections (e.g., bypass was opened). Among all samples from the pilot study of UV systems without prefiltration, no *E. coli* was detected (n=24). Total first-year costs for locally available UV systems with and without prefiltration ranged from $1,059-$1,645 and $927-$1,183, respectively, while operation and maintenance (O& M) costs ranged from $166-$266 and $142-$146, respectively. Given these data, UV systems may be a viable option for generating potable water; however, clear purchasing and installation protocols are needed as well as simple O&M guidelines for households to reduce user error.

**Synopsis:** Point-of-entry UV systems were able to produce water for domestic use with no detectable E. coli in 95.2% of samples among USVI households using roof-harvested rainwater.

## Introduction

Rain catchment systems are a common mechanism for supplementing household water supplies or accessing a primary water source across the Caribbean (Girona-Mata, 2020). In the US Virgin Islands (USVI), >90% of households have a rainwater catchment system consisting of roof-harvesting and collection of water in cisterns (often below ground and >20,000 liters of volume) via gutters and downspouts (Government of the United States Virgin Islands, 2018). Water is then distributed throughout the premise plumbing via a small pump and pressure tank. As the municipal water system reaches <25% of households (Government of the United States Virgin Islands, 2018), the majority of households rely on roof-harvested rainwater for domestic and potable needs. However, data from a 2019 evaluation of cistern water quality in USVI found that 80% of households using rain catchment systems had detectable levels of *Escherichia coli* (*E. coli*) in their cistern water (Consortium, 2020; Rao et al., 2022). Therefore, the availability of effective household water treatment options, point-of-entry (POE) or point-of-use (POU), is critical for supporting households in generating potable water.

Locally available water treatment options include both POE and POU systems leveraging UV light, filtration, or chlorine, while boiling and bottled water are also options utilized in USVI (Voth-Gaeddert et al., 2022a, 2022b). One POE water treatment option hypothesized to be effective at producing potable water for the entire house is the UV treatment system. These systems are installed immediately after the pump and pressure tank, treating the water before it moves to the remaining premise plumbing and taps. Most of the UV treatment systems are sold with one to two prefilters, ranging from 20µm to 1µm in pore size, to ensure turbidity is at manufacturer recommended levels for UV treatment (e.g., <1 NTUs). However, due to high capital costs, limited effectiveness data, and limited information provided to the public, these UV systems may be underutilized. Therefore, Love City Strong, Inc (LCS), a local non-governmental organization, facilitated a UV pilot program to provide enrolled households access to UV systems and support resources as well as to evaluate the effectiveness of the UV systems.

To evaluate the effectiveness of one brand of UV system available in USVI, two approaches were taken. First, longitudinal data from the UV pilot program was evaluated to assess the effectiveness of the UV system with two prefilters (5µm and 1µm in pore size) at producing tap water with no detectable levels of *E. coli*. Second, a follow-on pilot study was conducted to evaluate the effectiveness of the UV system without prefilters at producing tap water with no detectable levels of *E. coli*. Finally, capital and operation and maintenance (O&M) costs were estimated for a hypothetical household interested in purchasing a UV system.

## Methods

### Setting

USVI has a total population of 106,405 across three primary islands, St. Croix, St. Thomas, and St. John (United States Census Bureau, 2010). Rain-harvesting is the primary water source for the majority of households (Government of the United States Virgin Islands, 2018). As of 2020, mean monthly rainfall ranges from 36 mm in February to 142 mm in September (National Oceanic and Atmospheric Administration, 2021). This water is used for many household activities including cooking, cleaning, showering, brushing teeth, and drinking, among others (Government of the United States Virgin Islands, 2018). As of 2010, the median annual income for an individual is $24,704, and 32.5% of the population is below the poverty line (national mean is 15.1%) (United States Census Bureau, 2010).

### UV Access Program

In June 2018, LCS initiated a pilot program to provide UV systems and supplementary resources to households on St. John. Enrolled households were provided a Rainfresh Rainwater Filtration System (two prefilters and a UV bulb; model#: RW8, Richmond Hill, Ontario, Canada), free installation, training on maintenance, and free monthly testing for up to 12 months. The Rainfresh UV system operates at a flow rate of 30 LPM (8 GPM), provides a fluence (i.e., UV dose) of >40 mJ/cm^2^ (bulb has a 1-year lifespan), and has a recommended maximum turbidity level for the UV mechanism of <1 NTUs. Upon the initial household visit, a baseline survey was conducted collecting data on household and water quality characteristics. Two baseline water samples were collected; one from the cistern and one from the tap. Water quality parameters were tested onsite while a portion of the sample was placed on ice and sent to the laboratory on St. John for microbial contamination testing (see details below).

Next, the UV system was installed just after the pump and pressure tank by a certified plumber. Field teams would return to the household monthly to collect paired water samples from the cistern and a tap post-treatment (most often the kitchen tap, but some taps were available outside the household immediately post-treatment). The field team would also conduct an inspection of the UV system to ensure proper operating procedures were being followed. If the inspection or water testing indicated suboptimal effectiveness of the system, corrective action was taken. At the end of the free testing period, the household completed an exit survey and then assumed full responsibility for operating the UV system.

### UV without Prefiltration

As the prefiltration units can be a costly component of the UV system, a pilot study was conducted to evaluate the effectiveness of UV systems without prefiltration. The prefilter’s primary role is to reduce the turbidity of the water to acceptable operating conditions for the UV light to effectively transmit through the water and kill microbial contaminants. Previous data, and data from the UV program, suggest prefiltration may not be necessary to produce water with no detectable fecal indicator bacteria (Christensen and Linden, 2003; World Health Organization, 2017). Therefore, two volunteer households with the Rainfresh Rainwater Filtration System (using prefilters) enrolled in a pilot study. Three baseline samples were collected over a one-week period with the UV system including prefiltration in place following the same sampling procedures used in the UV system access program (described below). The prefilters were then removed, leaving the functional UV unit. Paired cistern and tap water samples were collected three times per week for four weeks. Upon completion of the four-week study, new prefilters were replaced in the UV system.

### Water Quality Parameters and Analyses

During each visit for both the UV access program and the UV pilot study, paired cistern and tap water samples were collected. For the UV access program, visits that occurred between June 2018 and July 2020 were included in the analysis (the program is ongoing). During the visit, a 500 mL polypropylene beaker was used to collect cistern water at a tap prior to treatment, after flushing the tap for 30 seconds (from here on referred to as the cistern sample). A separate beaker was used to collect a water sample from a tap, post-treatment, after flushing the tap for 30 seconds. The following water quality parameters were tested onsite immediately upon collection: pH, total dissolved solids (TDS), electrical conductivity (EC) (Hach Pocket Pro™+ Tester; Prod No. 9532800, Hach, Loveland, Colorado); free chlorine residual (FCR), total chlorine residual (TCR) (Hach Pocket Chlorimeter II; Prod No. 58700-00; LR-0.02); and turbidity (HACH 2100Q Portable Turbidimeter). In addition, 300 mL of each sample was transferred to a Whirl-Pak sample bag with a sodium thiosulfate tablet (Hach, Loveland, Colorado), placed on ice, and shipped to the laboratory on the island for processing within 12 hours. Microbial contamination was estimated by analyzing and quantifying total coliforms and *E. coli* via the IDEXX-Colilert-24® and Quanti-Tray/2000 (IDEXX Laboratories, Westbrook, Maine) using established methods (American Public Health Association (APHA), 2018).

To evaluate the effectiveness of the UV systems with prefilters and the UV systems without prefilters, the following analyses were conducted. First, descriptive statistics were generated for paired cistern-tap samples to evaluate overall pre-and post-installation characteristics of the water samples from the UV systems. Depending on the data, the mean, median, range, and standard deviation were estimated to compare pre-treatment and post-treatment samples (Stata Version 16 was used; (StataCorp LLC, 2019)). For the microbial data (total coliforms and *E. coli*), any values <1 MPN/100mL were set to 0.5 MPN/100mL while any values >2419.6 were set to 2420 MPN/100mL. All microbial data was then log10 transformed. Finally, capital and O&M costs of a set of locally available UV treatment systems were aggregated. This included the system used in the UV program along with other options available on the islands.

## Results

### UV Access Program: Descriptive Statistics

In total, data from N=51 households from the UV access program and N=271 household visits were evaluated to determine the effectiveness of the UV systems with prefiltration. Table 1 presents key descriptive statistics related to the households and associated cisterns. The baseline data suggest that the majority of households used concrete, underground cisterns, but not specifically for drinking water. The first household was enrolled in the program in June 2018 and the mean number of repeat visits for a household was 5.4.

**Table 1.** UV Filtration System Household Descriptive Statistics

| Measure | Category | n | Prevalence (%) \| Median (range) |
| --- | --- | --- | --- |
| Number of household members | *Median (range)* | 50 | 2<br>(1-10) |
| Drinking water | Bottled | 45 | 90% |
|  | Cistern | 5 | 10% |
| Number of cisterns | 1 | 27 | 54% |
|  | 2 | 21 | 42% |
|  | 3 | 2 | 4% |
| Age of cistern (years) | *Median (range)* | 43 | 34<br>(8-71) |
| Material of cistern | Concrete | 41 | 82% |
|  | Concrete/Plastic | 2 | 4% |
|  | Fiberglass | 2 | 4% |
|  | Plastic | 5 | 10% |
| Location of cistern | Above Ground | 20 | 41% |
|  | Underground | 29 | 59% |
| Cistern last cleaned (years) | *Median (range)* | 27 | 7<br>(1-12) |
|  | Never | 10 | 20.4% |
|  | Unknown | 12 | 24.5% |

*One household did not receive a baseline survey.*

### UV Access Program: Effectiveness of Water Treatment

Baseline samples were collected once for each household from the cistern (N=50) and the tap (N=51) and are presented in Table S1 and Figures S1a-e. Mean log10-MPN/100mL concentrations of total coliforms in the baseline cistern and tap samples were 2.68 and 2.18, respectively, while for *E. coli* concentrations were 1.06 and 0.30. Table 2 presents the results of the water quality parameters as well as concentrations of total coliforms and *E. coli* for all samples collected from cisterns and taps after installation of the UV systems with prefiltration.

**Table 2.** UV Access Program Cistern and Tap Sample Descriptive Statistics

| Measure | Cistern |  |  | Tap |  |  |
| --- | --- | --- | --- | --- | --- | --- |
|  | N | Median (range) | Mean (SD) | N | Median (range) | Mean (SD) |
| pH | 267 | 7.60<br>(4.21-10.7) | 7.70<br>(0.98) | 271 | 7.50<br>(3.75-10.9) | 7.54<br>(0.91) |
| Total Dissolved Solids (TDS) | 267 | 57.2<br>(8.00-582) | 95.9<br>(98.9) | 271 | 58.5<br>(10.3-621) | 101<br>(109) |
| Electric Conductivity ($\mu\text{S}/\text{cm}$) (EC) | 267 | 99.0<br>(7.56-854) | 165<br>(175) | 271 | 101<br>(14.5-1240) | 176<br>(196) |
| Free Chlorine (mg/L) | 249 | 0.02<br>(0.00->8.80) | 0.09<br>(0.59) | 254 | 0.02<br>(0.00-0.33) | 0.03<br>(0.04) |
| Total Chlorine (mg/L) | 250 | 0.03<br>(0.00->8.80) | 0.10<br>(0.59) | 253 | 0.02<br>(0.00-1.59) | 0.04<br>(0.11) |
| Turbidity (NTU) | 265 | 1.19<br>(0.00-18.7) | 2.21<br>(2.79) | 267 | 0.97<br>(0.00-7.43) | 1.42<br>(1.41) |
| Total Coliforms* | 263 | 2.51<br>(-0.30-3.38) | 2.01<br>(1.37) | 267 | -0.30<br>(-0.30-3.38) | 0.13<br>(0.96) |
| *E. coli* * | 263 | 0.72<br>(-0.30-3.38) | 0.92<br>(1.23) | 267 | -0.30<br>(-0.30-2.76) | -0.23<br>(0.38) |
| * Sample concentrations <1 and >2419.6 MPN/100 mL were assigned as 0.5 and 2420 MPN/100 mL, respectively, before log<sub>10</sub> transformation; 1 log = 10<sup>1</sup> MPN/100 mL |  |  |  |  |  |  |

Electrical conductivity at the tap after the installation of the UV system with prefiltration was higher than baseline tap samples while turbidity was reduced. Mean log10 MPN/100mL concentrations of total coliforms pre-(cistern) and post-(tap) treatment were 2.01 and 0.13, respectively, while mean *E. coli* concentrations were 0.92 and -0.23, respectively.

Table 3 and Figure S2a-b presents tap data on the occurrence of non-detections of total coliforms and *E. coli* after the UV systems with prefiltration. For all tap samples, including samples where the cistern had no contamination, *E. coli* was not detected in 95.2% (n=258) of tap samples (N=271) while total coliforms were not detected in 76.4% (n=207) of tap samples. For tap samples where the cistern sample had detectable levels of *E. coli* (N=171), *E. coli* was not detected in 93.6% (n=160) of paired tap samples, while for only those samples where the cistern sample had detectable levels of total coliforms (N=217), total coliforms were not detected in 72.4% (n=157) of paired tap samples. Table S2 presents data on the concentration of *E. coli* and total coliforms among tap samples with positive detections.

**Table 3.** Prevalence of Non-Detections for *E. coli* and Total Coliforms among Tap Samples (excluding baseline)

| Measure | n | Prevalence |
| --- | --- | --- |
| <1 *E. coli* | 258<br>(N=271) | 95.2% |
| Taps with <1 *E. coli* with cistern reading of $\geq 1$ *E. coli* | 160<br>(N=171) | 93.6% |
| <1 total coliforms | 207<br>(N=271) | 76.4% |
| <1 total coliforms with cistern reading of $\geq 1$ total coliforms | 157<br>(N=217) | 72.4% |

64.1% (171/267) of cistern samples had $\geq 1$ *E. coli*; 81.3% (217/267) of cistern samples had $\geq 1$ total coliforms

For all tap samples, post treatment, 13 had positive detections of *E. coli* while 64 had positive detections of total coliforms. As turbidity is an important water quality metric for effectiveness of UV systems, Table 4 presents the mean turbidity levels between the samples with positive *E. coli* or total coliform detections and the samples where *E. coli* or total coliforms were not detected. The data show turbidity values were higher among samples with no detection of *E. coli* suggesting turbidity may not have been the driver of detecting *E. coli* or total coliforms. In addition, of the 13 positive *E. coli* detections, two happened in repeated monthly visits of the same system suggesting course-correction/remediation of the issue was often possible by the field team (anecdotes of common reasons for user error-driven failure from field teams are provided in the discussion). For total coliforms, 64 positive detections occurred while 13 occurred in repeated monthly visits of the same system.

**Table 4.** Turbidity Readings Among Tap Samples (N=267)

| Measure | n | Turbidity<br>NTUs (mean, SD) |
| --- | --- | --- |
| Taps with $\geq 1$ *E. coli* | 13 | 1.3<br>(1.0) |
| Taps with $< 1$ *E. coli* | 253 | 1.5<br>(1.9) |
| Taps with $\geq 1$ *Total Coliforms* | 64 | 1.3<br>(1.2) |
| Taps with $< 1$ *Total Coliforms* | 202 | 1.6<br>(2.1) |

Four samples in Table 4 did not have turbidity data.

### UV without Prefiltration: Effectiveness of Water Treatment

To evaluate the necessity of the prefiltration system prior to the UV disinfection process, two systems were evaluated with only the UV system in place. Table 5 presents detailed data over the four-week trial period while Table S3 provides the full set of data (including baseline data). Baseline cistern water quality parameters were comparable to the systems evaluated in the UV system access program. Trial data suggested that no *E. coli* detections at the tap occurred for either system (N=24), while total coliforms were detected at the tap among six samples (25% or 75% non-detections) over both systems (N=24). However, only two total coliform detections were >2 MPN/100mL.

**Table 5.** UV System Without Prefiltration Trial Results

| Measure | Household 1 (N=12) |  | Household 2 (N=12) |  |
| --- | --- | --- | --- | --- |
|  | Cistern | Tap | Cistern | Tap |
| pH (mean, SD) | 7.59<br>(0.24) | 7.50<br>(0.35) | 7.67<br>(0.14) | 7.56<br>(0.16) |
| Total Dissolved Solids (TDS) (mean, SD) | 92.7<br>(19.1) | 128<br>(34.9) | 49.1<br>(10.6) | 47.6<br>(10.1) |
| Electric Conductivity (μS/cm) (EC) (mean, SD) | 153<br>(28.9) | 214<br>(57.0) | 74.5<br>(6.8) | 76.0<br>(5.3) |
| Free Chlorine (mg/L) (mean, SD) | 0.02<br>(0.03) | 0.02<br>(0.02) | 0.02<br>(0.03) | 0.03<br>(0.05) |
| Total Chlorine (mg/L) (mean, SD) | 0.03<br>(0.03) | 0.03<br>(0.03) | 0.04<br>(0.03) | 0.02<br>(0.02) |
| Turbidity (NTU) (mean, SD) | 0.55<br>(0.61) | 0.37<br>(0.51) | 1.28<br>(1.79) | 0.52<br>(0.67) |
| Total Coliforms* (mean, SD) | 1.81 | 0.10 | 3.31 | -0.28 |
|  | (0.42) | (0.72) | (0.24) | (0.09) |
| *E. coli* * (mean, SD) | 0.30<br>(0.48) | -0.30<br>(0.00) | 1.69<br>(0.67) | -0.30<br>(0.00) |
| * Sample concentrations <1 and >2419.6 MPN/100 mL were assigned as 0.5 and 2420 MPN/100 mL, respectively, before log<sub>10</sub> transformation; 1 log = 10<sup>1</sup> MPN/100 mL |  |  |  |  |

### Cost Analysis

Finally, the cost to the household for each system was estimated. These estimates may vary depending on the specific brand of UV system, plumber, and supply chain/procurement process (direct from company, via hardware store, NGO facilitated, etc.). Here we present data of the a range of local costs (from hardware stores and online) based on the following system requirements: 22.7-30.3 LPM (6-8 GPM) and fluence (i.e., UV dose) of 16-30 mJ/cm^2^. Table S4 presents the retail cost of the actual systems used in the program (however, bulk ordering agreements impacted the significance of this information). First year costs for a local UV system with and without prefiltration would total $1,059-$1,645 (with prefiltration) and $927-$1,183 (without prefiltration), while operation and maintenance (O&M) costs – including UV bulb and filter replacement – after the first year would total $166-$266 (with prefiltration) and $142-$146 (without prefiltration). Table 6 provides a cost breakdown for each system.

**Table 6.** Costs of the UV system with and without prefiltration

| Costs | UV with prefiltration** | UV without prefiltration** |
| --- | --- | --- |
| UV system | \$568-\$1099 | \$486-\$747 |
| Installation (additional parts, labor at \$100/hr) | Labor & Parts: \$400 | Labor & Parts: \$400 |
| Replacement parts (1 bulb, filters) | Bulb: \$60<br>2 Filters: \$24-\$120 | Bulb: \$60<br>No Filters |
| Energy costs (\$0.41/kW-hr) | \$79-\$86<br>22W-24W | \$79-\$86<br>22W-24W |
| **First Year Total\*** | \$1,059-\$1,645 | \$927-\$1,183 |
| **Annual Total** | \$166-\$266 | \$142-\$146 |

*\* only need to replace two filters the first year as unit comes with two filters initially; \*\* range includes three systems, the lower being PURA with UV system at 30.3 LPM (8 GPM) and 16mJ/cm<sup>2</sup> plus one prefilter and upper being Mighty Pure with UV system at 22.7 LPM (6 GPM) and 30mJ/cm<sup>2</sup> plus two prefilters. Viqua costs were between the cost of the two other brands.*

## Discussion

The data from the UV system access program suggested that *E. coli* was not detected in 95.2% of tap samples when the UV system with prefiltration was in place. This value only drops slightly when evaluating only those paired samples when detectable levels of *E. coli* were present in the cistern (93.6%). In addition, turbidity levels were lower among samples with detectable levels of *E. coli* (n=13) and total coliforms (n=64) compared to those without detectable *E. coli* or total coliforms. This suggests that the “failures” of the system may not have been related to turbidity issues. Field staff reported that for the majority of “failures,” a user error or wider system issue was observed. Examples include: a bypass valve around the system had been switched on, the UV system had been switched off or unplugged, insufficient electrical capacity triggering a breaker flip, or the UV system did not automatically restart after a power outage. This highlights the need for clear, non-technical guidance and information to accompany any type of water treatment technology when the operator will be a household member or non-expert.

In addition, total coliforms were not detected in 76.4% of all samples and 72.4% of tap samples where a positive detection was confirmed in the cistern. Of the 64 positive detections, 13 were repeat detections (back-to-back months). In comparison, for community water systems (CWS), the Environmental Protection Agency (EPA) states that to comply with the maximum contaminant level rules, no more than 5% of samples per month can have a positive detection for total coliforms (US Environmental Protection Agency, 2013). For USVI households, if a positive total coliform detection post-treatment co-occurs with a positive *E. coli* detection, resampling should be undertaken if possible and any obvious corrective action taken to the treatment system or associated plumbing. However, as the majority of samples were taken from the kitchen tap, the post-treatment premise plumbing may be a potential source for the higher number of total coliform detections due to biofilm-based microorganisms and lack of free chlorine residual (Falkinham et al., 2015; Proctor et al., 2022). Field staff were able to recall sampling locations for a small subset of samples both immediately post-treatment and at the kitchen tap. Of those paired samples where no detection of *E. coli* at the tap occurred and total coliforms were detected in the cistern sample, total coliforms were detected immediately post-treatment 17% of the time (n=5 of 29) compared to 31% of the time (n=11 of 35) among kitchen tap samples. Further evaluations should be conducted to understand the role of the premise plumbing in contributing to microbial contamination of household water as well as the potential for microbial regrowth post treatment.

In addition to the UV system with prefiltration, we evaluated the effectiveness of the UV system without prefiltration in two systems over a four-week period. The data suggested that out of N=24 samples, none had detectable levels of *E. coli* while the mean turbidity at the tap was 0.44 NTUs (range: 0.00-2.03 NTUs). As the sample size was small and the monitoring duration was short within this pilot study, further testing should be conducted. However, the data suggests that it may be feasible to maintain a sufficient level of microbial removal via the UV system without prefiltration, as this could save a household time and money. However, flow rate and fluence (UV dose) are also critical parameters for treatment efficacy in these systems especially without prefiltration. Incorporating a factor of safety in estimating a maximum or peak flow rate for a household (e.g., showering + washing machine + two faucets) can provide a locally adapted recommendation for what maximum flow rate to recommend in the UV systems. In aggregating local UV cost data, we used systems rated for 22.7 LPM (6 GPM) or greater. For fluence, the relationship between fluence, turbidity, flow rate, and microbial kill rate has been extensively explored previously but is often dependent on local source water characteristics including contamination risk and the type and size of turbidity causing material (Batch et al., 2004; Cantwell et al., 2008; Christensen and Linden, 2003; Craik, 2002; Liu, 2005; Liu and Zhang, 2006; Malley, 2000; Passantino et al., 2004; Severin et al., 1983). However, data suggests UV can be effective for turbidity levels up to 8-10 NTUs. The US EPA Surface Water Treatment Rule states that for small, unfiltered water systems the maximum allowable turbidity level is 5 NTUs (US Environmental Protection Agency, 2006) while the World Health Organization states that in low resource settings a goal of <5 NTUs should be maintained (World Health Organization, 2017). Given that measured turbidity levels in USVI cisterns are often well below 5 NTUs, partially due to the high residence time (for settling to occur), turbidity may not be a significant concern. However, limited data currently exists on chemical contamination in USVI cistern water (e.g., nitrates, lead, arsenic, etc.) which may influence prefiltration aspects of water treatment.

The fluence level (UV dose) of the system evaluated in this study was >40 mJ/cm^2^ while other locally available UV systems may provide a fluence as low as 30 or 16 mJ/cm^2^. While the National Sanitation Foundation/American National Standards Institute (NSF/ANSI) 55 standards designate UV systems with a fluence >40 mJ/cm^2^ as class A and >16 mJ/cm^2^ as class B, most pathogens are inactivated at 5-10 mJ/cm^2^ (Malayeri et al., 2016). However, the type of microbial contaminant can have a significant influence on the effectiveness of the UV system (Malayeri et al., 2016). In this study, we only used fecal indicator bacteria (total coliforms and *E. coli*). Microorganisms more resistant to UV light (e.g., Adenovirus) may be present (Malayeri et al., 2016) and therefore a system with a higher fluence should be used when possible. If a UV system with a low fluence is used (e.g., 16 mJ/cm^2^), households should ensure the total flow rate at a single point-in-time is as low as possible. Given previous literature and local data, households should, at a minimum, use UV systems rated for 22.7-30.3 LPM (6-8 GPM) and generate a fluence of 16-30 mJ/cm^2^. However, if a household experiences high flow rates, turbidity issues, or are concerned about sewage contamination in their cistern, a higher fluence UV system should be used (e.g., 40mJ/cm^2^). A UV system with a NFS/ANSI 55 Class A certification has been extensively tested and provides a minimum fluence of 40mJ/cm^2^. Further field research should be conducted to evaluate the field effectiveness in USVI for lower rated UV systems and for pathogen-specific targets.

A central attribute to water treatment access is cost and local availability. Therefore, we evaluated the costs associated with the UV systems (with and without prefiltration) available to households on the islands. UV system specifications included a flow rate between 22.7-30.3 LPM (6-8 GPM) and a fluence of between 16-30 mJ/cm^2^. The data suggested that households with limited available finances, could access a local UV system without prefiltration, including installation, for $927-$1,183 for the first year and an O&M cost of $142-$146. For a UV system with prefiltration, these costs increase to $1,059-$1,645 and $166-$266, respectively. There was a significant cost increase between single and double prefiltration options. In addition, the cost of installation was almost as much as some of the UV systems themselves (i.e., ∼$400 vs $486).

Comparing the costs of the UV systems to other possible point-of-entry (POE) and point-of-use (POU) water treatment systems available on the islands is difficult given the varying efficacies of the treatment systems and the locations within the house in which these treatment systems provide treated water (i.e., all taps, one tap, no taps). For example, an under the sink, reverse osmosis system may cost ∼$300-400 for the unit (plumbing not included), but only provide treated water at the sink. A counter-top ceramic filter is also ∼$300-400 for the unit (no plumbing necessary), but the household must continue to fill and use this water from its designated location. Both systems can provide potable water (Berkey Filters, 2022). For whole-house treatment (POE), fewer options are available. Some households use only membrane filtration (∼$200-400), but depending on pore size, at best this can protect against large protozoan-based cysts (≤1 micron absolute filter), while bacteria and viruses can easily pass through. Finally, passive chlorination has been demonstrated as a viable technique to provide potable water (Lindmark et al., 2022) and low-cost design options are currently being developed and tested for use in USVI (Voth-Gaeddert et al., 2022a). These systems may cost $150-200 for the units once fully developed and operational. Therefore, given the variety of water use habits, water treatment technology costs and efficacies, and stratification of socio-economic levels in USVI, decision support tools can be an important tool to help facilitate data driven decisions by households in accessing water treatment technologies.

Finally, given this evaluation was based on a field program, several limitations are present in this study. First, only one type of treatment system was evaluated in the study; however, the structural dynamics and system specifications of the system reflect other systems available on island. We recommend further evaluations be conducted on lower fluence and flow rate systems to evaluate effectiveness. In addition, the outcome indicators utilized in this study were total coliforms and *E. coli* which are common fecal indicator bacteria. However, certain pathogens are more resistant to UV light and therefore a higher fluence should be used when possible. Next, not all UV system units at households were sampled the same number of times which could cause some units to be overrepresented in the aggregated statistics. In addition, post-treatment sampling points were not systematically recorded and ranged from immediately post-treatment to the kitchen tap. Finally, the pilot study on the UV system without prefiltration only included two systems over a one-month period (N=12 samples each). Further research over a wider range of fluence and turbidity levels would help support the cost-saving option of not using prefiltration. Despite these limitations, the data presented here provide an important contribution to understanding the effectiveness of water treatment options available in USVI.

## Conclusion

In this study, we evaluated data from a local program providing access to UV systems with prefiltration and data from a pilot study on the effectiveness of the same UV systems without prefiltration. *E. coli* was not detected in 95.2% of samples (N=271) collected from taps with UV systems with prefiltration, while no *E. coli* was detected in samples (N=24) collected from UV systems without prefiltration. Turbidity was not related to the probability of detecting *E. coli* in the UV systems. The first-year costs of the UV systems ranged from $927-$1,183 for UV systems without prefiltration to $1,059-$1,645 for UV systems with prefiltration. O&M costs ranged from $142-$146 to $166-$266, respectively. Finally, robust and easy to use guidelines should be developed for households to ensure proper operation and minimize user error. UV systems can offer an effective option for water treatment at the whole-house level in USVI while decision support tools and user guidelines will be important to empowering households to make data-driven decisions.

## Data Availability

All data produced in the present study are available upon reasonable request to the authors

## Acknowledgements

The authors are grateful for the dedication of the Love City Strong staff for their support and commitment to the people of the US Virgin Islands.

## Author Contribution Statement

**Lee Voth-Gaeddert**: Conceptualization, Methodology, Data curation, Supervision, Writing-Original draft preparation. **Douglas Momberg**: Data curation, Formal analysis, Visualization, Writing-Reviewing and Editing. **Kela Brathwaite**: Conceptualization, Investigation, Writing-Review and Editing. **Andrew Schranck**: Conceptualization, Writing-Review and Editing. **Mandy Lemley**: Conceptualization, Investigation, Project administration, Writing-Review and Editing. **Stephen Libbey**: Conceptualization, Project administration, Resources, Writing-Review and Editing.

