## Supplementary Material for "Evaluating the Effectiveness of Point-of-Entry UV Treatment for Cistern Water Among Households in the US Virgin Islands"

**Supplementary Materials**

Voth-Gaeddert et al. "*Household UV Treatment of Cistern Water as a Viable Option for Generating Potable Water in the US Virgin Islands*"

**Table S1.** UV Filtration System Baseline Cistern and Tap Descriptive Statistics

| **Measure** | **Cistern** (N = 50) | | **Tap** (N = 51) | |
|---|---|---|---|---|
| | Median (range) | Mean (SD) | Median (range) | Mean (SD) |
| pH (mean, SD) | - | 7.6 (0.87) | - | 7.7 (0.82) |
| Total Dissolved Solids (TDS) | 67.9 (10-312) | 89.5 (70.3) | 71 (13-649) | 103.3 (104.4) |
| Electric Conductivity (µS/cm) | 112.8 (20-622) | 162.4 (137.8) | 115 (26-1300) | 105.7 (205.2) |
| Free Chlorine (mg/L) | 0.02** (0->8.8) | 0.03 (0.03) | 0.02 (0->8.8) | 0.05 (0.09) |
| Total Chlorine | 0.02** (0->8.8) | 0.03 (0.03) | 0.03 (0->8.8) | 0.04 (0.08) |
| Turbidity (NTU) | 0.9 (0.01-5.43) | 1.07 (0.9) | 0.85** (0-57) | 2.8 (8.6) |
| Total Coliforms* | 2.68 (-0.30-3.38) | 2.3 (1.2) | 2.18 (-0.30-3.38) | 1.8 (1.3) |
| *E. coli** | 1.06 (-0.30-3.38) | 1.2 (1.2) | 0.30 (-0.30-3.30) | 0.8 (1.2) |

* Samples concentrations <1 and >2419.6 MPN/100 mL were assigned as 0.5 and 2420 MPN/100 mL, respectively, before log10 transformation; 1 log = $10^1$ MPN/100 mL
** N = 49

**Figure S1a.** Box plot of baseline pH values.

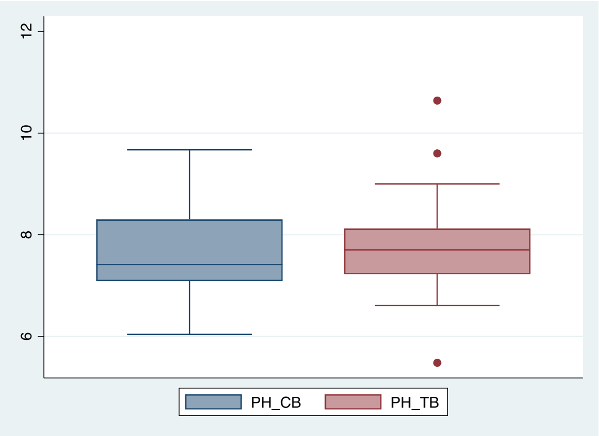

*CB = cistern baseline (N=50); TB = tap baseline (N=51)*

**Figure S1b.** Box plot of baseline TDS values.

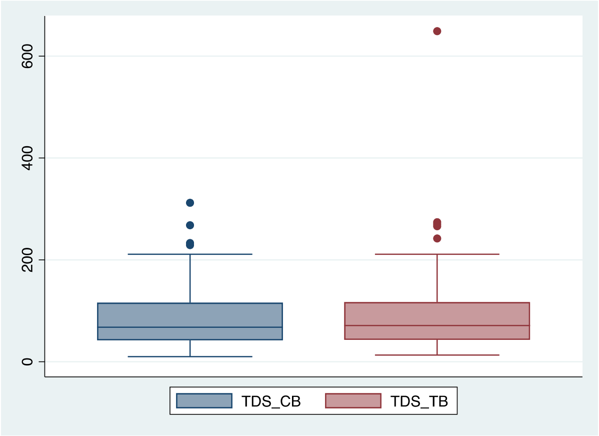

*CB = cistern baseline (N=50); TB = tap baseline (N=51)*

**Figure S1c.** Box plot of baseline electrical conductivity values.

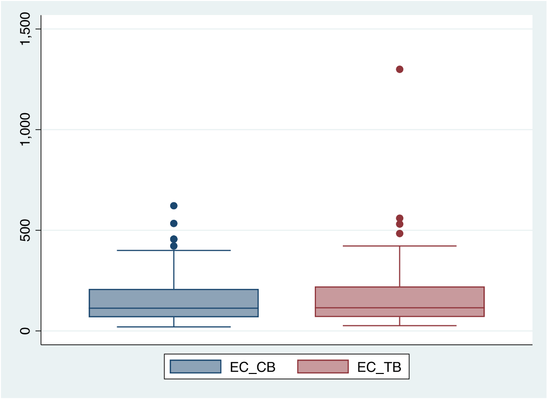

*CB = cistern baseline (N=50); TB = tap baseline (N=51)*

**Figure S1d.** Box plot of baseline free chlorine values (ppm)

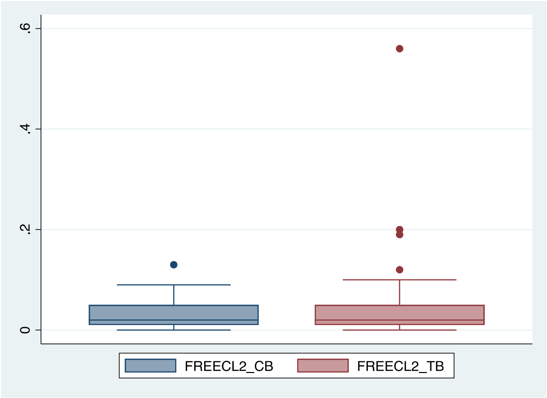

*CB = cistern baseline (N=49); TB = tap baseline (N=51)*

**Figure S1e.** Box plot of baseline total chlorine values (ppm)

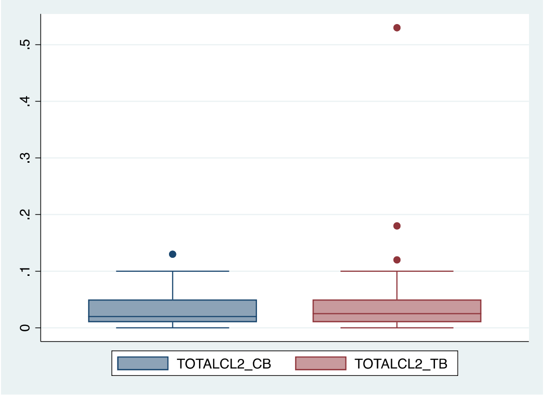

*CB = cistern baseline (N=50); TB = tap baseline (N=51)*

**Figure S1f.** Box plot of baseline total coliform values.

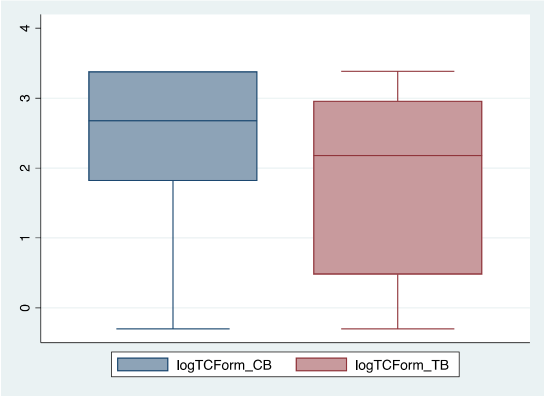

*CB = cistern baseline (N=50); TB = tap baseline (N=51)*

**Figure S1e**. Box plot of baseline *E. coli* values.

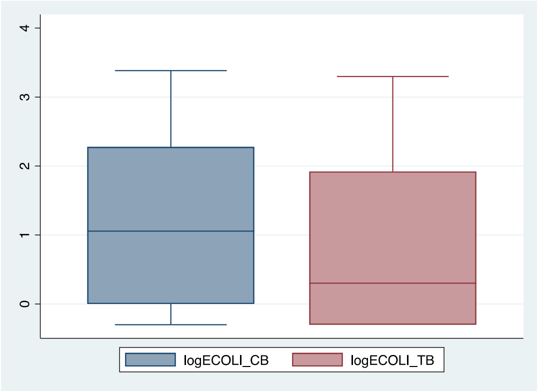

*CB = cistern baseline (N=50); TB = tap baseline (N=51)*

**Figure S2a.** Box plot of total coliform values pre and post UV treatment (N=263)

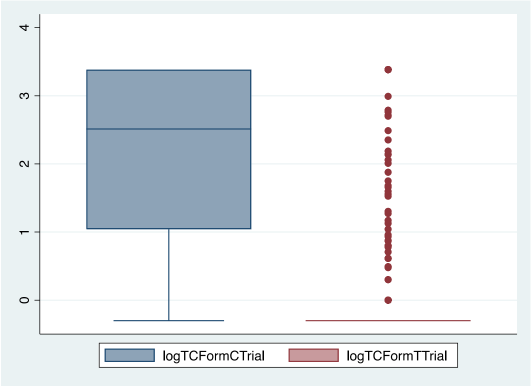

**Figure S2b.** Box plot of *E. coli* values pre and post UV treatment (N=263)

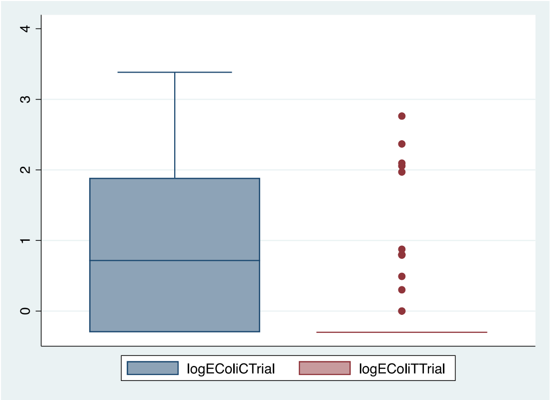

**Table S2.** Concentration of *E. coli* and Total Coliforms Among Positive Detections in Tap Samples (excluding baseline; N=271)

| **Measure** | **Positive Detections** | **MPN/100mL** Mean (SD) | **Log10 MPN/ 100mL** Mean (SD) |
|---|---|---|---|
| ≥1 *E. coli* | 4.8% (n=13) | 90.6 (157) | 1.96 (2.20) |
| ≥1 total coliforms | 23.6% (n=64) | 492 (903) | 2.69 (2.96) |

**Table S3**. Full water quality data on two UV-only systems for baseline and trial.

| **Measure** | **Household 1** | | | | **Household 2** | | | |
|---|---|---|---|---|---|---|---|---|
| | **Cistern** | | **Tap** | | **Cistern** | | **Tap** | |
| | **Baseline** | **Trial** | **Baseline** | **Trial** | **Baseline** | **Trial** | **Baseline** | **Trial** |
| | **N=3** | **N=12** | **N=3** | **N=12** | **N=3** | **N=12** | **N=3** | **N=12** |
| pH (mean, SD) | 7.8 (0.2) | 7.6 (0.2) | 7.8 (0.1) | 7.5 (0.4) | 7.7 (0.1) | 7.7 (0.1) | 7.7 (0.1) | 7.6 (0.2) |
| Total Dissolved Solids (TDS) (mean, SD) | 84.5 (8.7) | 92.7 (19.1) | 83.8 (16.8) | 127.9 (34.9) | 38.6 (1.0) | 49.1 (10.6) | 42.0 (5.7) | 47.6 (10.1) |
| Electric Conductivity (μS/cm) (EC) (mean, SD) | 145.1 (11.9) | 153.4 (28.9) | 138.8 (27.8) | 213.5 (57.0) | 64.5 (1.9) | 74.5 (6.8) | 69.4 (9.2) | 76 (5.3) |
| Free Chlorine (mg/L) (mean, SD) | 0.0 (0.0) | 0.0 (0.0) | 0.0 (0.0) | 0.0 (0.0) | 0.0 (0.0) | 0.0 (0.0) | 0.0 (0.0) | 0.0 (0.1) |
| Total Chlorine (mg/L) (mean, SD) | 0.0 (0.0) | 0.0 (0.0) | 0.0 (0.0) | 0.0 (0.0) | 0.2 (0.2) | 0.0 (0.0) | 0.0 (0.0) | 0.0 (0.0) |
| Turbidity (NTU) (mean, SD) | 0.6 (0.2) | 0.6 (0.6) | 0.8 (0.4) | 0.4 (0.5) | 2.0 (1.2) | 1.3 (1.8) | 0.3 (0.3) | 0.5 (0.7) |
| Total Coliforms* (mean, SD) | 1.6 (0.5) | 1.8 (0.4) | -0.2 (0.2) | 0.1 (0.7) | 3.4 (0.0) | 3.3 (0.2) | 0.1 (0.6) | -0.3 (0.1) |
| *E. coli** (mean, SD) | 1.2 (0.5) | 0.3 (0.5) | -0.3 (0.0) | -0.3 (0.0) | 1.3 (0.2) | 1.7 (0.7) | -0.3 (0.0) | -0.3 (0.0) |
| * Samples concentrations <1 and >2419.6 MPN/100 mL were assigned as 0.5 and 2420 MPN/100 mL, respectively, before log10 transformation; 1 log = $10^1$ MPN/100 mL | | | | | | | | |

**Table S4**. Costs of the UV system used in the program (actual costs)

| **Costs** | **UV with prefiltration (program)** | **UV without prefiltration (program)** |
|---|---|---|
| UV system | $1189 | $1189^ |

| | | |
|---|---|---|
| Installation (additional parts, labor at $100/hour) | Parts: $150<br>Labor: $400 | Parts: $100<br>Labor: $400 |
| Replacement parts (1 bulb, filters) | Bulb: $110<br>4 Filters: $212 | Bulb: $110<br>No Filters |
| Energy costs ($0.41/kW-hr) | $144<br>@40W | $144<br>@40W |
| **First Year Total**** | $1,989 | $1,883 |
| **Annual Total** | $466 | $254 |

Due to the nature of the program, the UV systems were procured at wholesale, but this does not provide a useful benchmark for households or field practitioners.
